# Ischemic stroke in patients with atrial fibrillation and low cardiovascular risk profile: A nationwide, registry-based cohort study

**DOI:** 10.1101/2025.04.25.25326461

**Authors:** Alexia Karagianni, Carmen Basic, Tatiana Zverkova Sandström, Aidin Rawshani, Zacharias Mandalenakis, Per Ladenvall

## Abstract

**Background:** Although there is substantial evidence supporting the prevention of ischemic stroke (IS) in high-risk patients with atrial fibrillation (AF), knowledge regarding AF patients with a low cardiovascular risk profile remains limited. Furthermore, the necessity of anticoagulant therapy in this population has been widely debated.

**Methods:** Data from Swedish health registers were utilized to identify all patients diagnosed with AF but without prior cardiovascular conditions between 1987 and 2018. The risk of IS was assessed using Cox regression models, with AF patients compared to age- and sex-matched controls without AF.

**Results:** The study included a total of 229,075 patients with AF and 455,541 matched controls without AF. The overall risk of IS was 2.3 times higher (95% confidence interval [CI] 2.2–2.3) in patients with AF compared to controls over a mean follow-up period of 9 years (range 3–32 years). Women with AF had a 4.4-fold increased risk of developing IS within the first year following diagnosis (hazard ratio [HR] 4.4, CI 4.2–4.7) compared to matched women without AF. Additionally, younger patients with AF (aged 35–49 years) exhibited the highest risk of IS within the first year after diagnosis (HR 8.3, 95% CI 4.0– 17.1).

**Conclusions:** This large, nationwide, register-based cohort study found that even in the absence of cardiovascular conditions, patients with atrial fibrillation had more than double the risk of ischemic stroke compared to matched controls. The risk was especially elevated in women and younger individuals, particularly within the first year after AF diagnosis. These findings underscore the urgent need to refine risk stratification and explore preventive strategies beyond traditional clinical risk factors.

## Introduction

Atrial fibrillation (AF) is the most prevalent arrhythmia encountered in clinical practice and is associated with significant morbidity, notably increasing the overall risk of ischemic stroke (IS) (1–4). Clinical risk scores have been developed as tools to identify AF patients who may benefit from oral anticoagulation therapy. According to current European guidelines for the management of AF, gender has been excluded as a risk stratification parameter in clinical risk scoring, leading to the adoption of the CHA_2_DS_2_-VA score instead of the CHA_2_DS_2_-VASc score (2). However, existing studies indicate that women are frequently underdiagnosed with respect to atrial arrhythmias, which subsequently may lead to undertreatment and an elevated risk of IS (5,6).

Additionally, non-valvular AF patients with comorbidities beyond those accounted for in the CHA_2_DS_2_-VA score, such as renal failure and congenital heart disease, have been shown to exhibit an increased risk of IS (7–13). The occurrence of AF among younger individuals is frequently associated with a range of contributing factors, including obesity, insufficient or excessive physical activity, tobacco and alcohol use, sleep-disordered breathing, and psychosocial determinants (14). Additionally, existing literature indicates that comorbid conditions such as malignancies and chronic pulmonary diseases may function as additional risk stratifiers, further elevating the risk of ischemic stroke in patients with AF (15–19).

Threrefore, the efficacy and appropriateness of anticoagulation therapy for the prevention of IS in patients with AF and non-cardiovascular conditions remain subjects of ongoing investigation and debate, particularly within younger patient populations (20–25).

Given the rising incidence of IS in younger individuals (26–28), there is an urgent need for more robust evidence regarding the risk of IS in AF patients without conventional cardiovascular conditions. The present study aims to investigate the incidence of IS among AF patients without cardiovascular conditions and to evaluate their relative risk of developing IS in comparison to matched controls without AF.

## Methods

### Data source

A unique, personal identification number is given to every permanent resident in Sweden and it is obligatory to use it for all authorities and healthcare systems. Thus, it is possible to follow-up each individual over time through Swedish health registers. The Swedish National Patient Register provides information of all hospitalizations nationwide with a complete cover of all hospitals in Sweden since 1987. The Swedish National Patient Register provides also information of all hospital outpatient visits since 2001. The Cause of Death Register carries detailed information about major and contributing causes of death since 1961. Validation studies have shown that both Swedish National Patient and the Cause of Death Register provide data with high quality, and they widely used for epidemiological studies (29–31). The Swedish Prescribed Drug Register (SPDR) was established in July 2005 and includes data on all prescribed drugs purchased at pharmacies in Sweden, containing actual information of the drug utilization and it has been validated as high quality register (32).

### Definitions

In Sweden, each diagnosis is coded according to the International Statistical Classification of Diseases (ICD) and Related Health Problems (ICD 8^th^ revision until 1986, ICD 9^th^ revision until 1996, ICD 10^th^ revision thereafter). The definitions of atrial fibrillation, ischemic stroke (IS), cardiovascular conditions (such as heart failure, diabetes mellitus, ischemic heart disease, stroke/TIA or peripheral arterial disease), and the other comorbidities according to the ICD-8, ICD-9, and ICD-10 codes, are summarized in supplementary Table 1 A. The Anatomical Therapeutic Chemical classification (ATC) codes of anticoagulant agents are summarized in supplementary Table 1 B.

The study baseline was defined as the date of the AF onset. Therefore, the baseline characteristics refer to these dates. The primary endpoint of the study was ischemic stroke (IS). No distinction was made among paroxysmal, persistent or permanent AF.

### Study Population

We linked data from the Swedish National Patient Register and the Cause of Death Register in order to identify all patients with AF diagnosis in Sweden from January 1^st^, 1987 to December 31^st^, 2018. For each patient with AF, two controls were selected from the Swedish Total Population Register, matched for birth year, sex and residence. The follow-up of the study was from January 1^st^, 1987 until December 31^st^ 2018. The baseline was the time of AF diagnosis.

We excluded all individuals with a previous cardiovascular condition (such as heart failure, diabetes mellitus, ischemic heart disease, stroke/TIA or peripheral arterial disease), renal failure, congenital heart disease and valvular heart disease, at the time of AF diagnosis.

Patients with other comorbidities not related to cardiovascular conditions known to elevate the risk of atrial fibrillation (AF), such as chronic obstructive pulmonary disease (COPD) and cancer, were identified both at baseline and during follow-up. However, individuals who developed IS prior to their diagnosis of COPD or cancer were excluded from the sub-analysis.

By using the SPDR register which introduced in July 2005, we identified patients on anticoagulant treatment at baseline (2006) and at follow-up (Supplemental Table 2).

Patients with AF and their matched controls were divided into six age groups based on age at baseline.

### Ethical considerations

The study complied with the Declaration of Helsinki, and the study protocol was approved by the Regional Ethical Review Board in Gothenburg, Sweden.

### Statistics

All statistical analyses were performed using SAS 9.4 software. The matching was made by age, sex and index year after excluded patients with pre-existing cardiovascular conditions. Continuous variables are expressed as mean ± standard deviation (SD) and median (interquartile range(IQR)), and categorical variables are expressed as count and percentage. The incidence rate of ischemic stroke per 1,000 person-years was estimated for patients with AF and controls . A Cox regression model using univariate analysis was used to estimate the hazard ratios (HRs) and 95% CIs. The time scale in the model was the date of atrial fibrillation diagnosis intervention until 31 December 2018 or until primary end point or death. The cumulative incidence of ischemic stroke is presented for the patients with AF and controls without AF. In a sub-analysis, we estimated the cumulative incidence of stroke in the cohort between 2006 to 2017 that were under anticoagulant treatment. A P-value of <0.05 was considered statistically significant.

## Results

In total 229,075 patients with AF and 455,541 matched controls without AF were included in the present study. The characteristics for the study population are shown in the Table 1. The mean follow-up of the study was 8.7 years (SD 6.8) and women represented the 44.4 % of the study population. The majority of the patients with AF and controls (76%) was above the age of 60 years.

**Table 1.**
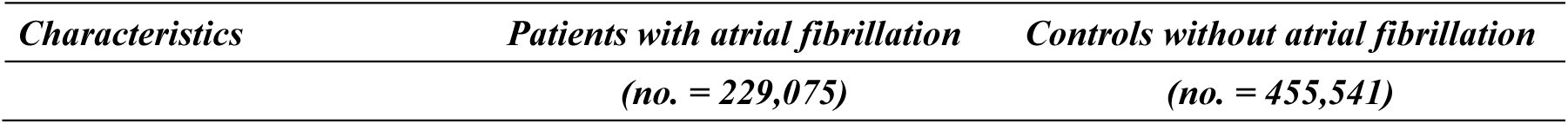
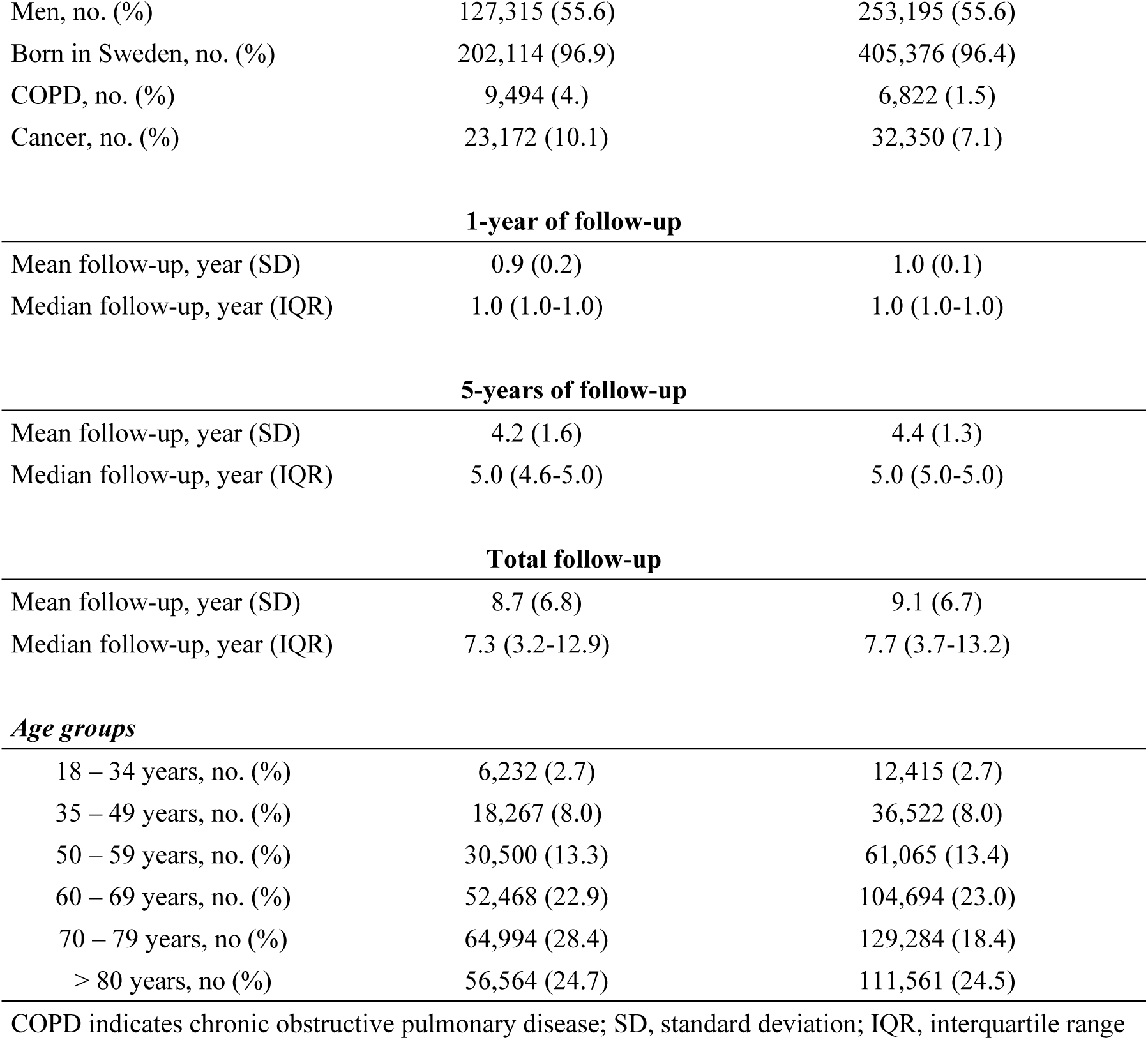
Characteristics of the study population.

| *Characteristics* | *Patients with atrial fibrillation* | *Controls without atrial fibrillation* |
| --- | --- | --- |
|  | *(no. = 229,075)* | *(no. = 455,541)* |
| Men, no. (%) | 127,315 (55.6) | 253,195 (55.6) |
| Born in Sweden, no. (%) | 202,114 (96.9) | 405,376 (96.4) |
| COPD, no. (%) | 9,494 (4.) | 6,822 (1.5) |
| Cancer, no. (%) | 23,172 (10.1) | 32,350 (7.1) |
| **1-year of follow-up** |  |  |
| Mean follow-up, year (SD) | 0.9 (0.2) | 1.0 (0.1) |
| Median follow-up, year (IQR) | 1.0 (1.0-1.0) | 1.0 (1.0-1.0) |
| **5-years of follow-up** |  |  |
| Mean follow-up, year (SD) | 4.2 (1.6) | 4.4 (1.3) |
| Median follow-up, year (IQR) | 5.0 (4.6-5.0) | 5.0 (5.0-5.0) |
| **Total follow-up** |  |  |
| Mean follow-up, year (SD) | 8.7 (6.8) | 9.1 (6.7) |
| Median follow-up, year (IQR) | 7.3 (3.2-12.9) | 7.7 (3.7-13.2) |
| **Age groups** |  |  |
| 18 – 34 years, no. (%) | 6,232 (2.7) | 12,415 (2.7) |
| 35 – 49 years, no. (%) | 18,267 (8.0) | 36,522 (8.0) |
| 50 – 59 years, no. (%) | 30,500 (13.3) | 61,065 (13.4) |
| 60 – 69 years, no. (%) | 52,468 (22.9) | 104,694 (23.0) |
| 70 – 79 years, no. (%) | 64,994 (28.4) | 129,284 (18.4) |
| > 80 years, no. (%) | 56,564 (24.7) | 111,561 (24.5) |

COPD indicates chronic obstructive pulmonary disease; SD, standard deviation; IQR, interquartile range

In total, 30,095 patients with AF (13.14 %) and 28,644 controls (6.29 %) developed IS. The incidence rates and the risk of IS during 1-year, 5-year and total period of the study among patients with AF and controls are shown in Table 2. The overall risk of IS was 2.3 times higher (95% confidence interval [CI] 2.2–2.3) in patients with AF compared to controls (Figure 1).

**Figure 1.**
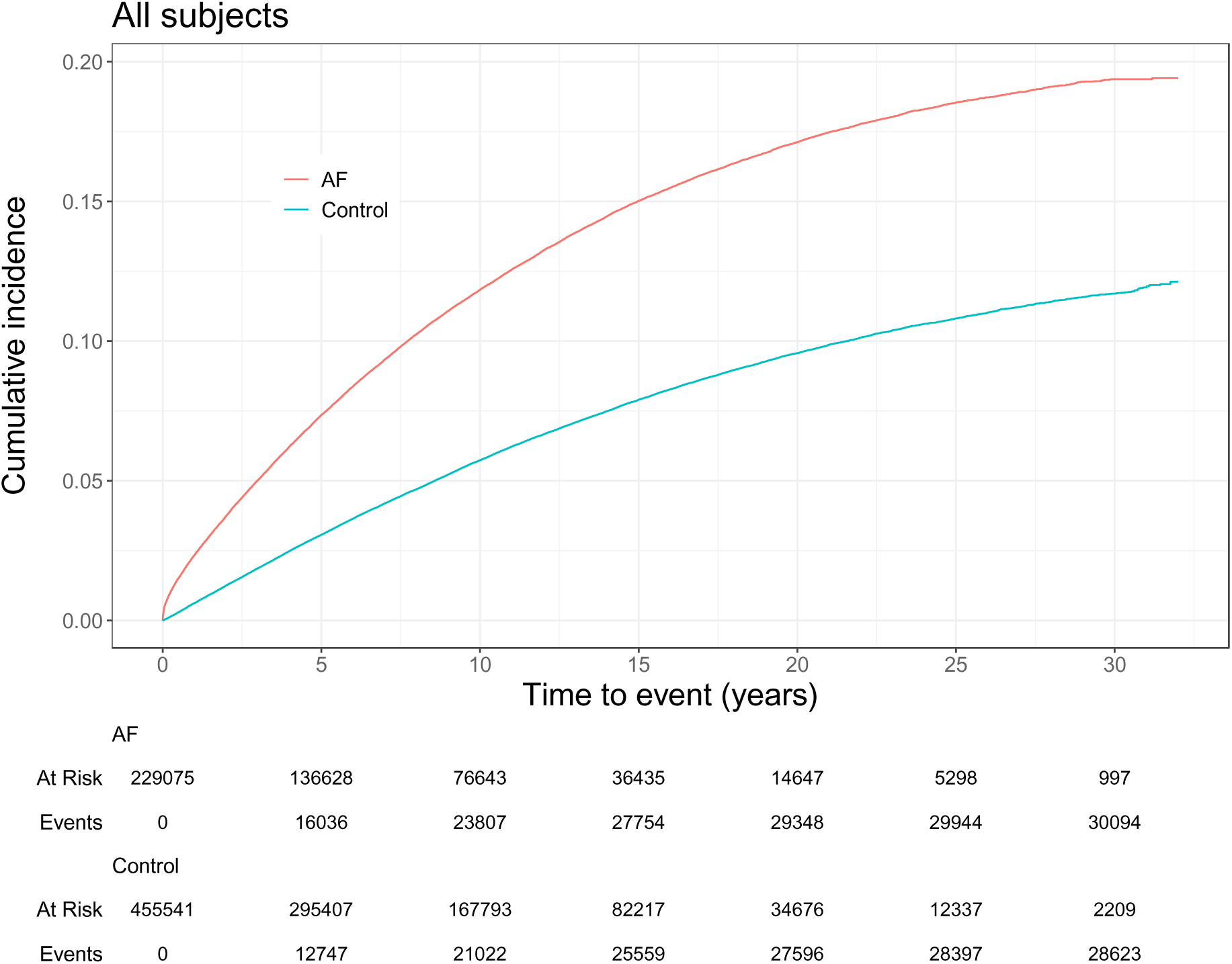
Cumulative incidence of ischemic stroke in the study population

**Table 2.**
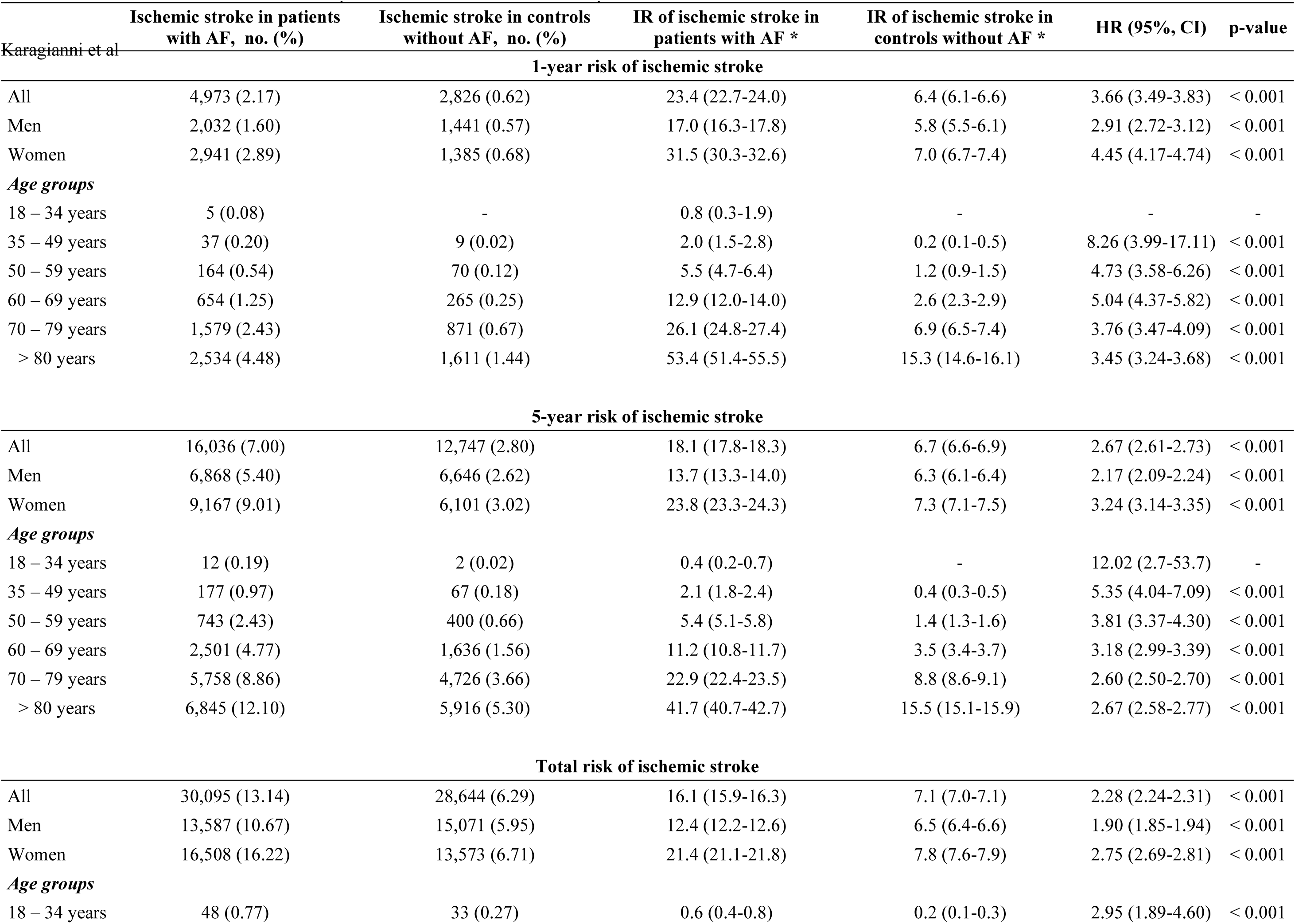
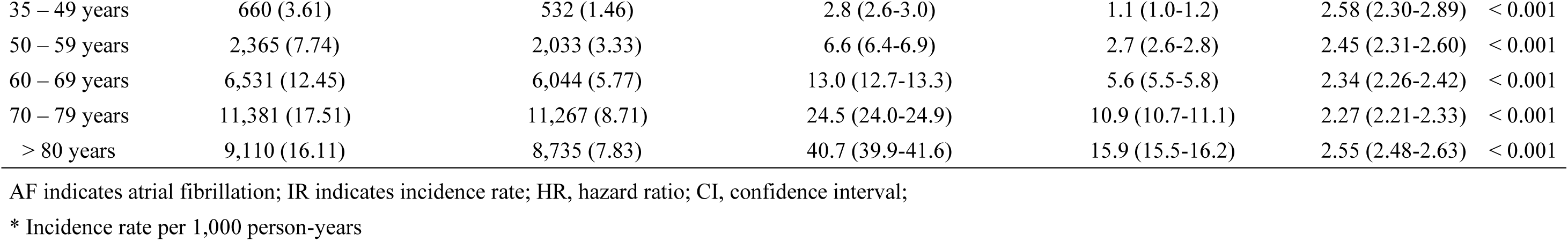
Risk of ischemic stroke in low-risk patients with atrial fibrillation compared to matched controls.

| Karagianni et al | Ischemic stroke in patients<br>with AF, no. (%) | Ischemic stroke in controls<br>without AF, no. (%) | IR of ischemic stroke in<br>patients with AF * | IR of ischemic stroke in<br>controls without AF * | HR (95%, CI) | p-value |
| --- | --- | --- | --- | --- | --- | --- |
| **1-year risk of ischemic stroke** |  |  |  |  |  |  |
| All | 4,973 (2.17) | 2,826 (0.62) | 23.4 (22.7-24.0) | 6.4 (6.1-6.6) | 3.66 (3.49-3.83) | < 0.001 |
| Men | 2,032 (1.60) | 1,441 (0.57) | 17.0 (16.3-17.8) | 5.8 (5.5-6.1) | 2.91 (2.72-3.12) | < 0.001 |
| Women | 2,941 (2.89) | 1,385 (0.68) | 31.5 (30.3-32.6) | 7.0 (6.7-7.4) | 4.45 (4.17-4.74) | < 0.001 |
| **Age groups** |  |  |  |  |  |  |
| 18 – 34 years | 5 (0.08) | - | 0.8 (0.3-1.9) | - | - | - |
| 35 – 49 years | 37 (0.20) | 9 (0.02) | 2.0 (1.5-2.8) | 0.2 (0.1-0.5) | 8.26 (3.99-17.11) | < 0.001 |
| 50 – 59 years | 164 (0.54) | 70 (0.12) | 5.5 (4.7-6.4) | 1.2 (0.9-1.5) | 4.73 (3.58-6.26) | < 0.001 |
| 60 – 69 years | 654 (1.25) | 265 (0.25) | 12.9 (12.0-14.0) | 2.6 (2.3-2.9) | 5.04 (4.37-5.82) | < 0.001 |
| 70 – 79 years | 1,579 (2.43) | 871 (0.67) | 26.1 (24.8-27.4) | 6.9 (6.5-7.4) | 3.76 (3.47-4.09) | < 0.001 |
| > 80 years | 2,534 (4.48) | 1,611 (1.44) | 53.4 (51.4-55.5) | 15.3 (14.6-16.1) | 3.45 (3.24-3.68) | < 0.001 |
| **5-year risk of ischemic stroke** |  |  |  |  |  |  |
| All | 16,036 (7.00) | 12,747 (2.80) | 18.1 (17.8-18.3) | 6.7 (6.6-6.9) | 2.67 (2.61-2.73) | < 0.001 |
| Men | 6,868 (5.40) | 6,646 (2.62) | 13.7 (13.3-14.0) | 6.3 (6.1-6.4) | 2.17 (2.09-2.24) | < 0.001 |
| Women | 9,167 (9.01) | 6,101 (3.02) | 23.8 (23.3-24.3) | 7.3 (7.1-7.5) | 3.24 (3.14-3.35) | < 0.001 |
| **Age groups** |  |  |  |  |  |  |
| 18 – 34 years | 12 (0.19) | 2 (0.02) | 0.4 (0.2-0.7) | - | 12.02 (2.7-53.7) | - |
| 35 – 49 years | 177 (0.97) | 67 (0.18) | 2.1 (1.8-2.4) | 0.4 (0.3-0.5) | 5.35 (4.04-7.09) | < 0.001 |
| 50 – 59 years | 743 (2.43) | 400 (0.66) | 5.4 (5.1-5.8) | 1.4 (1.3-1.6) | 3.81 (3.37-4.30) | < 0.001 |
| 60 – 69 years | 2,501 (4.77) | 1,636 (1.56) | 11.2 (10.8-11.7) | 3.5 (3.4-3.7) | 3.18 (2.99-3.39) | < 0.001 |
| 70 – 79 years | 5,758 (8.86) | 4,726 (3.66) | 22.9 (22.4-23.5) | 8.8 (8.6-9.1) | 2.60 (2.50-2.70) | < 0.001 |
| > 80 years | 6,845 (12.10) | 5,916 (5.30) | 41.7 (40.7-42.7) | 15.5 (15.1-15.9) | 2.67 (2.58-2.77) | < 0.001 |
| **Total risk of ischemic stroke** |  |  |  |  |  |  |
| All | 30,095 (13.14) | 28,644 (6.29) | 16.1 (15.9-16.3) | 7.1 (7.0-7.1) | 2.28 (2.24-2.31) | < 0.001 |
| Men | 13,587 (10.67) | 15,071 (5.95) | 12.4 (12.2-12.6) | 6.5 (6.4-6.6) | 1.90 (1.85-1.94) | < 0.001 |
| Women | 16,508 (16.22) | 13,573 (6.71) | 21.4 (21.1-21.8) | 7.8 (7.6-7.9) | 2.75 (2.69-2.81) | < 0.001 |
| **Age groups** |  |  |  |  |  |  |
| 18 – 34 years | 48 (0.77) | 33 (0.27) | 0.6 (0.4-0.8) | 0.2 (0.1-0.3) | 2.95 (1.89-4.60) | < 0.001 |
| 35 – 49 years | 660 (3.61) | 532 (1.46) | 2.8 (2.6-3.0) | 1.1 (1.0-1.2) | 2.58 (2.30-2.89) | < 0.001 |
| 50 – 59 years | 2,365 (7.74) | 2,033 (3.33) | 6.6 (6.4-6.9) | 2.7 (2.6-2.8) | 2.45 (2.31-2.60) | < 0.001 |
| 60 – 69 years | 6,531 (12.45) | 6,044 (5.77) | 13.0 (12.7-13.3) | 5.6 (5.5-5.8) | 2.34 (2.26-2.42) | < 0.001 |
| 70 – 79 years | 11,381 (17.51) | 11,267 (8.71) | 24.5 (24.0-24.9) | 10.9 (10.7-11.1) | 2.27 (2.21-2.33) | < 0.001 |
| > 80 years | 9,110 (16.11) | 8,735 (7.83) | 40.7 (39.9-41.6) | 15.9 (15.5-16.2) | 2.55 (2.48-2.63) | < 0.001 |

AF indicates atrial fibrillation; IR indicates incidence rate; HR, hazard ratio; CI, confidence interval;
\* Incidence rate per 1,000 person-years

Analyses by age and sex demonstrated that the incidence of IS was higher in women compared to men over all ages group (HR women 2.75 (2.69-2.81) vs men 1.90 (1.85-1.94)) especially the first year after the diagnosis of AF (Table 2, Figure 2). The overall risk for IS was two times higher in men with AF compared to controls and similar to all age-groups (Supplemental Table 3). However, when compared to controls the risk for IS was higher in middle-aged women (35-59 years old) with AF compared to elderly (> 70 years old) women (Supplemental Table 4 and supplemental figure). Of note, the hazard ratio of IS between cases and controls was higher in young patients (< 50 years old) with AF compared to middle and older aged (Table 2). The cumulative incidence of IS of the study population according to age is presented in Figure 3.

**Figure 2.**
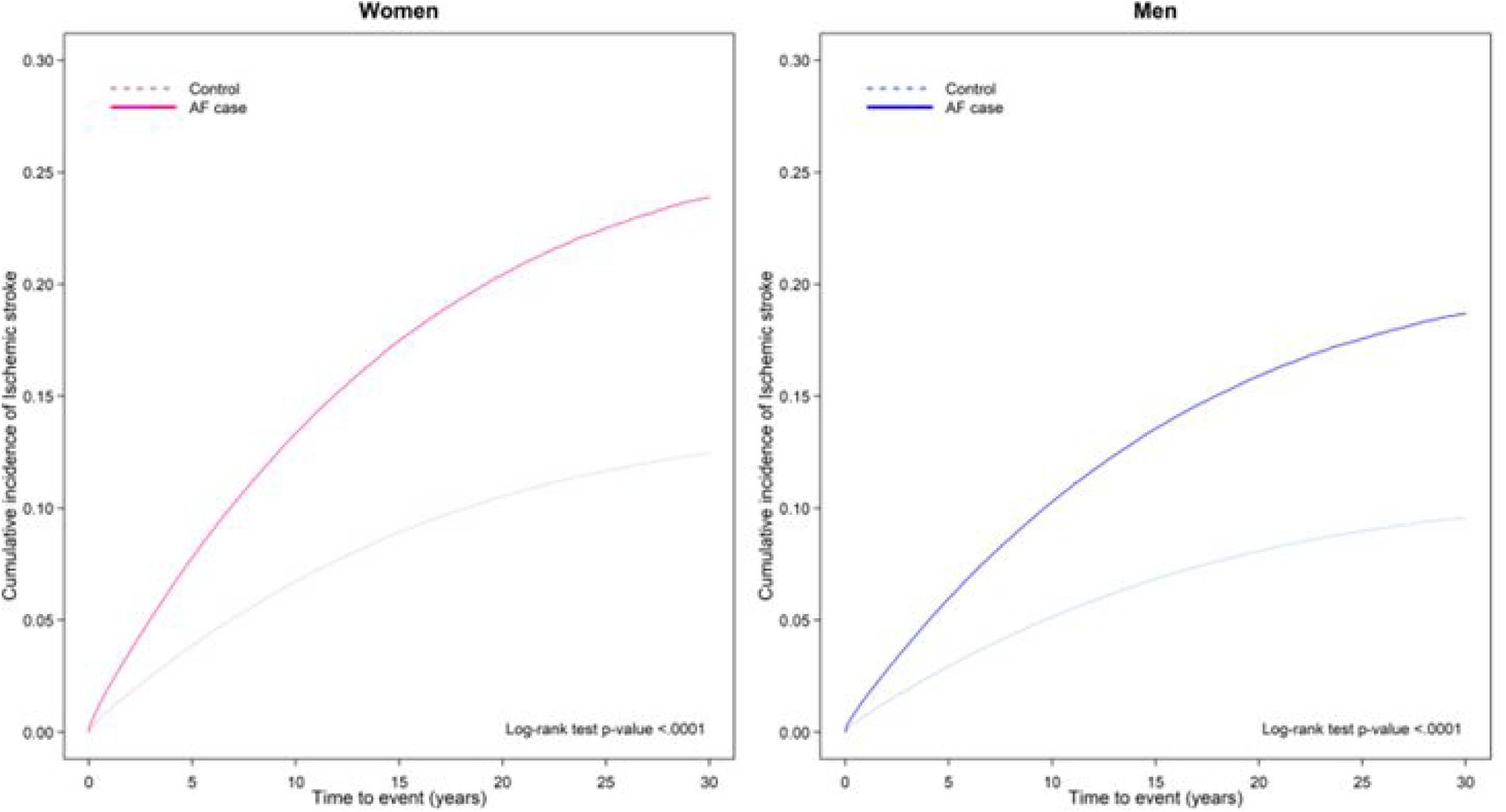
Cumulative incidence of ischemic stroke in the study population according to sex

**Figure 3.**
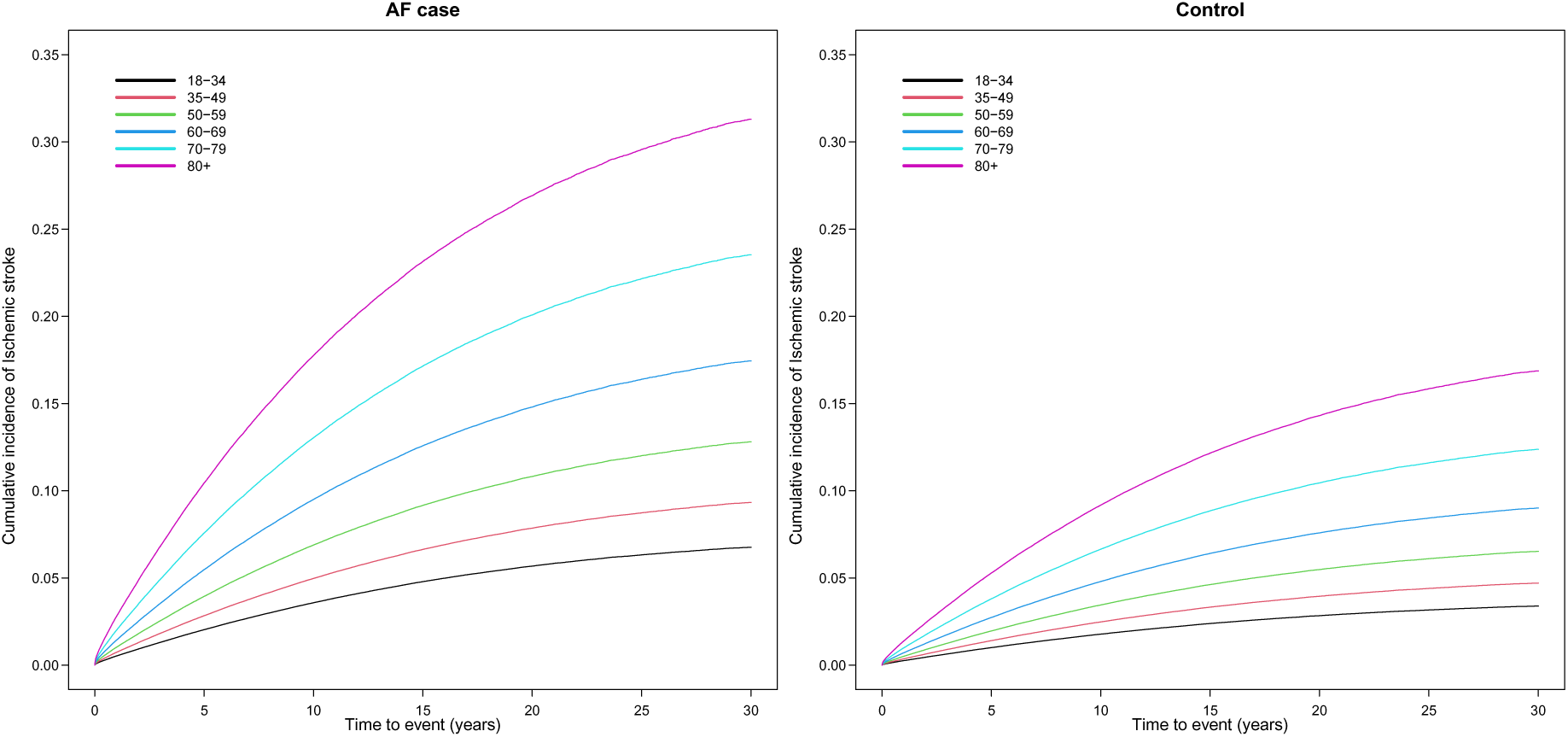
Cumulative incidence of IS of the study population according to age

Anticoagulation treatment increased during follow-up. At baseline, 39% of patients received anticoagulant treatment which increased to 78% during follow-up, while 14.6% of controls were treated at baseline which increased to 27% of controls during follow-up. Women had a higher use of anticoagulant treatment as expected (80% vs 29% women compared to men with atrial fibrillation). The incidence of stroke in patients on anticoagulant treatment was twice as high in patients with atrial fibrillation compared to controls, equally distributed in all age-groups (Table 3).

**Table 3.**
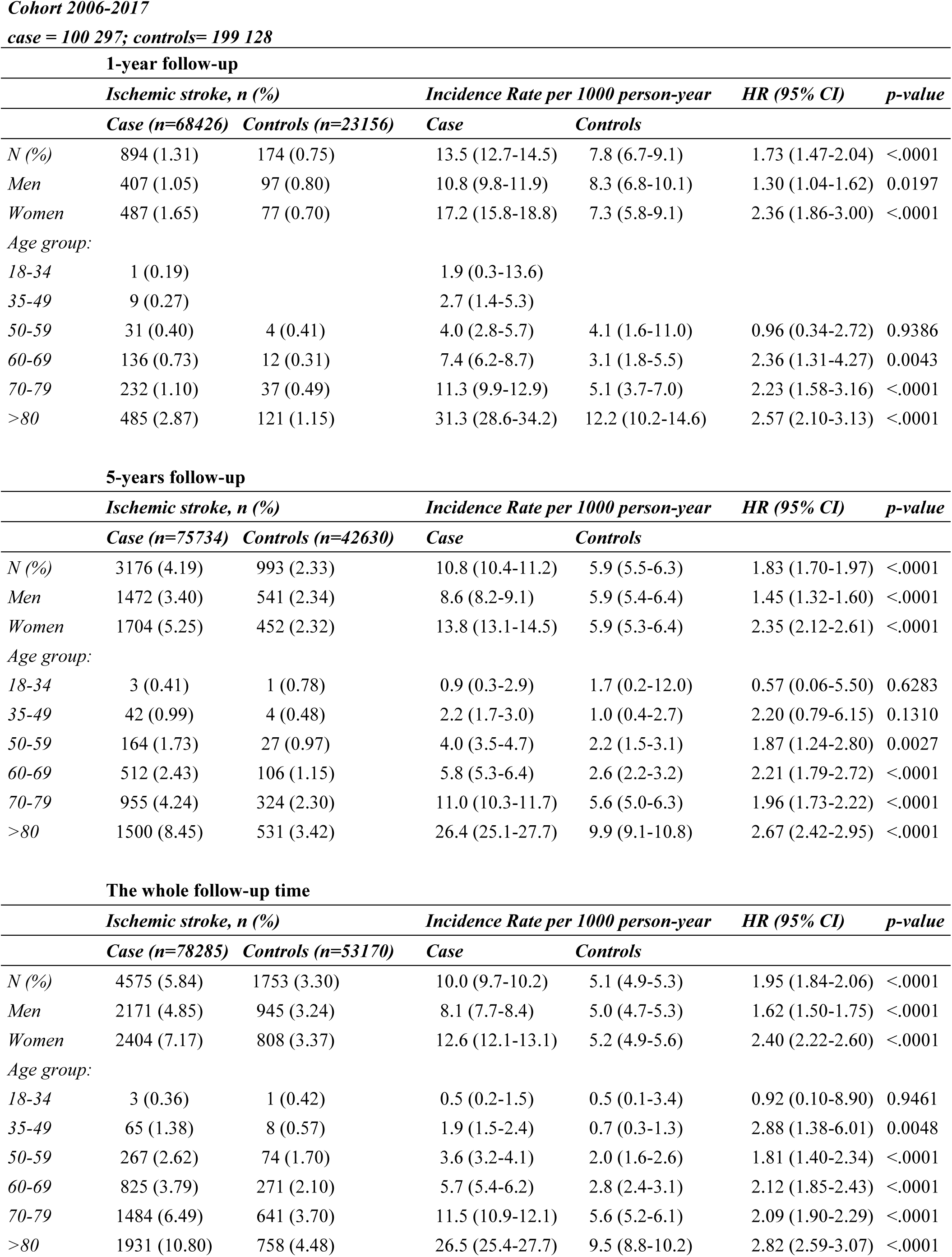
Incident ischemic stroke in patients and controls on anticoagulant treatment.

COPD and cancer incidence was almost 3-fold more frequent in patients with AF compared to controls and equally distributed between men and women (Supplemental Table 5). The HR for IS within the first year of diagnosis of AF in cancer young and middle-aged patients was 5.8, CI 1.26-26.97 and 4.9, CI 2.93-8.31 (Supplemental Table 5A). The overall risk of IS in AF patients with COPD remained higher over two times fold during the whole follow-up time (Supplemental Table 5B).

## Discussion

In this large registry-based study, we observed that the incidence of IS among patients with AF and a low-risk cardiovascular profile was more than twice as high as that of age- and sex-matched controls without AF. Notably, this risk was most pronounced within the first year following AF diagnosis, after which it declined across all groups but remained elevated throughout the study period.

Our findings indicate that individuals aged 35–49 years faced an over eightfold increased risk of IS compared to controls during the first year following AF diagnosis, despite the absolute number of IS events being lower in this age group relative to older cohorts. Recent studies indicate a rising incidence of ischemic stroke (IS) among young adults over the past decades. Currently, IS in young adults accounts for approximately one-third of all IS cases, highlighting the ongoing uncertainty regarding optimal management strategies for this population, as they are often underrepresented in clinical trials (21). Notably, the cumulative risk of recurrent stroke can reach up to 15% within a 10-year period. While atherosclerotic risk factors are the primary contributors to stroke in older adults, younger individuals are affected by a wider range of etiological factors (33). Although the absolute number of strokes in this age group remains relatively low, IS in young adults has considerable clinical and socioeconomic implications, exerting a disproportionate impact on both society and the economy. In our study, young adults with atrial fibrillation (AF), even in the absence of cardiovascular conditions, exhibited a significantly elevated risk of IS within the first year following diagnosis compared to their counterparts without AF or cardiovascular conditions. One possible explanation is that AF itself serves as an independent risk factor for IS, or alternatively, that young adults diagnosed with AF may have additional, unaccounted-for risk factors not captured in current risk stratification models. Importantly, the risk of IS was also significantly elevated in middle-aged individuals (50–59 years) and those aged 60–69 years, who exhibited a fivefold greater risk of IS compared to controls of the same age and sex, with this risk surpassing that observed in older groups. A plausible explanation is that elderly patients with AF are more likely to be prescribed anticoagulants due to underlying comorbidities reflected in their CHA_2_DS_2_-VA scores. Notably, patients and control subjects were matched in terms of age and absence of cardiovascular conditions at baseline, underscoring the substantial role AF as an independent risk factor for IS.

Interestingly, our study revealed that the risk of IS stabilized across all age groups over the follow-up period, suggesting that the most substantial increase in risk occurs within the first year following AF diagnosis.

Recent European guidelines have removed sex as a factor in IS risk stratification for patients with AF (2). However, the influence of sex remains a topic of debate. A recent cohort study by Champsi et al. found that sex did not significantly affect the contemporary risk of adverse events, including IS, in patients with AF (34). Nonetheless, women experience a higher incidence of fatal strokes compared to men, and in the United States, stroke was the third leading cause of death among women compared to the fifth leading cause among men in 2019 (35). Our study similarly found that women with AF had a higher risk of IS compared to men, particularly within the first year following AF diagnosis, despite a greater proportion of women receiving anticoagulant therapy. The previous CHA_2_DS_2_-VASc score included female sex as a risk factor for IS, which may account for the higher proportion of women receiving anticoagulants.

The incidence of IS among patients with AF receiving anticoagulation was twice as high in those aged 60 years and above during the first year following AF diagnosis. Elderly patients generally have a higher prevalence of comorbidities, contributing to a greater incidence of IS despite anticoagulation therapy. Notably, the Swedish Prescribed Drug Register (SPDR) provides data on actual medication utilization, reducing potential bias related to nonadherence. Nevertheless, the overall risk of IS remained highest in the 35–49-year-old age group. Specifically, within this cohort, the risk of IS was three times higher in AF patients receiving anticoagulation compared to controls, whereas in older age groups, this relative risk was approximately twofold. One possible explanation is that younger anticoagulated patients may have additional comorbidities or complexities that necessitated early anticoagulation initiation.

In recent years, significant advancements in neuroimaging have substantially improved the diagnosis of IS. Modern imaging techniques, such as computed tomography (CT) and magnetic resonance imaging (MRI), have enhanced the accuracy of stroke syndrome identification and facilitated the determination of underlying etiologies. Consequently, there has been an increase in the detection of IS events, including subclinical cases that may have previously remained undiagnosed. However, despite advances in imaging techniques in recent years, our key observation from this study remains: younger patients with newly diagnosed atrial fibrillation exhibit a distinct complexity, with the highest risk of ischemic stroke occurring within the first year—risks not attributable to conventional stroke risk factors, as such conditions were excluded in the studied population.

Simultaneously, the widespread adoption of novel oral anticoagulants has been associated with a reduction in IS risk, particularly among patients with AF(36). These agents have demonstrated efficacy in decreasing the incidence of ischemic events. However, it remains challenging to ascertain whether this decline can be attributed exclusively to anticoagulants, as concurrent improvements in the management of other risk factors likely contribute to this trend. Additionally, the use of anticoagulation therapy is inherently associated with an elevated risk of bleeding, underscoring the need for careful patient selection and ongoing monitoring to optimize therapeutic outcomes (37).

In our study, we excluded patients with AF who had valvular disease, congenital heart disease, prior cardiovascular comorbidities, or renal failure, as these conditions are independently associated with increased IS risk (2,7–14). However, recent studies suggest that extracardiovascular conditions, such as COPD and cancer, may also serve as risk stratifiers for IS in patients with AF. Our findings align with previous research indicating a higher incidence of COPD and cancer among patients with AF, with IS incidence remaining elevated in these populations. Advances in cancer treatment have transformed it into a more manageable condition; however, both cancer and its treatment can impact cardiovascular health. The question of whether patients with cancer and AF should receive anticoagulation remains complex, with ongoing studies exploring this issue. Achieving a balance between preventing ischemic stroke (IS) with anticoagulation and minimizing the risk of bleeding is particularly challenging in patients with cancer, as they are at a significantly higher risk of bleeding complications compared to the population without cancer. Additionally, patients with COPD exhibit a higher prevalence of AF and an increased risk of IS, yet current guidelines do not provide clear recommendations regarding early anticoagulation in this population.

### Limitations and Strengths

This study utilized registry-based data, introducing the potential for diagnostic misclassification of atrial fibrillation and ischemic stroke.

Lifestyle-related factors—such as physical activity, sleep patterns, smoking, and alcohol consumption— as well as body mass index, were not captured in this study.

Moreover, we did not differentiate between paroxysmal, persistent, and chronic AF, which may introduce bias, particularly among younger patients. However, current AF management guidelines do not distinguish between these AF subtypes in terms of treatment strategy.

Additionally, the SPDR was established in July 2005; thus, patients who initiated anticoagulation therapy before this date were not included in our analysis.

However, this study included a large, nationwide cohort of over 229,000 patients with atrial fibrillation and nearly 456,000 matched controls, providing high statistical power and robust estimates. With a mean follow-up of 9 years and a maximum of 32 years, the study captures long-term outcomes and trends over time.

Data were derived from well-established Swedish national health registers with high diagnostic validity (85–95%), ensuring reliability of the findings. By restricting the cohort to individuals without prior conventional comorbidities and risk factors for IS, the study isolates the impact of atrial fibrillation itself on stroke risk, which is less well understood.

The study highlights a significantly increased stroke risk in so-called ’low-risk’ AF patients, particularly among younger individuals and women—groups that are often under-recognized in risk stratification.

### Conclusions

This large nationwide cohort study demonstrated that despite the low absolute risk of IS in young patients with AF and no cardiovascular conditions, their relative risk of IS was eightfold higher within the first year following AF diagnosis compared to age- and sex-matched controls. Furthermore, women exhibited a greater risk of IS, particularly within the first year following AF diagnosis, compared to men. These findings highlight the need for further research to assess the potential benefits of anticoagulation therapy in younger adults, particularly during the first year post-diagnosis, and to refine current clinical risk scores to incorporate additional risk stratifiers.

## Data Availability

The data that support the findings of this study are available from the corresponding author upon reasonable request.

## Disclosures

Per Ladenvall is an employee of AstraZeneca Biopharmaceuticals Research and Development, Gothenburg, Sweden.

The other authors declare no disclosure.

## Sources of Funding

This work was supported by Trygg-Hansas Forksningsstiftelse, the Emelie Fond, and the Swedish Research Council (2019-00193 SIMSAM).

